# Pediatric Device Clinical Trials Activity Compared to Drugs and Biologics Trials 1999-2022

**DOI:** 10.1101/2023.08.07.23293770

**Authors:** Joshua Dienstman, Stanley J. Stachelek, Abba M. Krieger, Kolaleh Eskandanian, Juan C. Espinoza, Michael R. Harrison, Chester J. Koh, Vasum Peiris, Erika Torjusen, Robert J. Levy

## Abstract

**Objective:** This study assessed the state of PMD development by comparing PMD clinical trials to pediatric trials evaluating drugs and biologics, from 1999-2022. We hypothesized changes in numbers and types of PMD trials compared to drugs and biologics represent an indicator of PMD growth.

**Study Design:** www.clinicaltrials.gov was used to identify and quantify both PMD clinical trials and pediatric trials for drugs and biologics. Clinical specialty was also assessed. The institutions included were the seven children’s hospitals primarily affiliated with the FDA PDC grants program between 2018-2023.

**Results:** 243 PMD clinical trials were identified based on the year of initiation. The average number of PMD trials initiated per year per institution was 1.5. PMD trials significantly increased (p=0.0083) from 2014 onward compared to pediatric clinical trials for drugs and biologics, which demonstrated no significant change in trial initiation activity. A more than five-fold increase in PMD trials was observed from 2014-2018 compared to previous time periods, and there were 48% more PMD trials from 2019-2022 compared to 2014-2018. PMD trials represented 5% of clinical trials at the institutions studied.

**Conclusions:** While clinical trial activity for drug and biologic development remained stable from 1999-2022, initiation of PMD trials significantly increased. The present results suggest that clinical trials growth reflects increased PMD development. Accommodation and promotion of PMD clinical trial activity, which is still relatively small, by relevant programs and policies at the institutional and government levels may foster the advancement of PMD to further address unmet needs.

**Article Summary:** This article is an analysis of device trials performed at seven children’s hospitals affiliated with the FDA Consortia grants program between 1999 and 2022.

**What’s Known on This Subject:** There have been no prior studies of device trial activity at a cohort of children’s hospitals at academic medical centers. Over the past decade, FDA programs have been initiated to assist stakeholders in advancing the development of pediatric medical devices.

**What This Study Adds:** Pediatric device trials account for only 5% of total trials at the institutions studied. Of note, only half of these PMD trials (2.4% of total clinical trials) were sponsored by industry and likely seeking pediatric labeling.

## Introduction

The lack of progress in development and commercialization of PMD is indicative of an important unaddressed healthcare inequity.^1–3^ PMD are used in all aspects of pediatric healthcare, including treating patients with rare diseases, or those at high risk.^4–7^ However, PMD development has in general lagged compared to adult devices. ^1, 3, 8^ This has been due to factors including business concerns and product liability issues with perceived higher risks for trials including children.^2, 9–11^ As a result, there is disproportionate use of many medical devices evaluated, authorized and intended for adults but used in children, exposing children to an unclear benefit-risk profile.^1, 12, 13^

The FDA PDC grants program, and the Program for Pediatrics and Special Populations (P&SP) at FDA’s CDRH, support select academic institutions in their efforts to advance PMD development. The Pediatric Medical Device Safety and Improvement Act of 2007 ^14, 15^ enabled the FDA to launch the PDC grants program in 2009 ^16^, and at present there are five PDC nationwide that are associated with seven primarily affiliated children’s hospitals. The PDC offer a number of services to support and encourage the development of PMD, and these include: sponsored project funding for PMD development, consultant services to assist with regulatory guidance and assessing intellectual property, and optimizing FDA interactions with PMD innovators. The PDC affiliated institutions, which are all academic children’s hospitals with both well-established clinical specialty expertise and extensive research infrastructure, have had a close association with the FDA PDC program and the strategic initiatives advanced by CDRH’s P&SP. Thus, the rationale for the present study was based on examining PMD clinical trials at these children’s hospitals because of their select interest in PMD development and unique interaction with FDA’s CDRH P&SP, and OOPD.

The present study analyzed PMD clinical trials data from 1999 to 2022 at the seven primary PDC children’s hospitals participating in the FDA PDC grant program period 2018-2023, and compared this activity to clinical trials for drugs and biologics between 1999 to 2022 at the same children’s hospitals. It was hypothesized that changes in numbers and types of PMD trials compared to drugs and biologics represent an indicator of PMD growth. Thus, the objective of this investigation was to identify and compare rates of growth, within the time period studied, with comparisons between the number of clinical trials focused on PMD versus those involving biologics or drugs. Secondary objectives were to identify clinical specialties that had increased PMD trial activity, and to identify temporal trends in the number of associated PMD clinical trials. We utilized the database resources and search functions of www.clinicaltrials.gov to carry out the relevant queries for this study.

## Methods

### Clinical trial sites studied

The PMD clinical trial locations of interest for this study were the primary seven children’s hospitals affiliated with the FDA’s PDC grants program: CHLA, CHOP, CNH, LPCHS, TCH, UCSF BCH, and UPMC CHP.

### Search methodology

A search for PMD trials was conducted on www.clinicaltrials.gov. The search was conducted using the following parameters as allowed by www.clinicaltrials.gov: Years, 1999 until 2022 (search closure date, 6/20/2022); Study Type: Interventional Studies (Clinical Trials); Recruitment Status: Not yet recruiting, recruiting, enrolling by invitation, active - not recruiting, and completed; Age Group: Child^17^ (birth-17 years old, ages 18-21 were not included because of search engine parameter restrictions); Intervention/Treatment: “device,”; Funding type: All (NIH, Industry, Other); and Location terms: Hospital Name/City. Trials that were suspended, terminated, withdrawn, or had status unknown were not included. All other available parameters were left at the default settings. For a comparison that reflects institutional clinical trial infrastructure, the same search criteria (changing the Intervention/Treatment search terms to “drug” and “biologic”) were used to search for drug and biologic trials at the institutions included in this study.

The search results were used to create databases for both medical device and drug-biologic trials, and these were used to review and verify data and remove incorrectly coded trials for each hospital. These data were reviewed for verification of the trial data, coding, and location to ensure all iterations of site names were included. The www.clinicaltrials.gov database does not include a parameter for device classification. Thus, the authors reviewed each institution’s trial data set to identify, per FDA device definition, specific device-related entries.^18, 19^ Clinical specialty was also determined by review of the device and trial description. It should be noted that the “Digital” specialty designation includes devices and applications that specifically fall within the FDA’s scope for device software functions and mobile medical applications. ^20, 21^ Also, the “Behavioral” specialty designation includes devices that address conditions relating to mental and psychological health such as mental illnesses and emotional disorders. This differentiates these devices from the Neurology/Neurosurgery specialty which includes devices that address conditions relating to the anatomy, functions, and organic disorders of the central and peripheral nervous system. The “Other” category designation includes devices that could not be classified by any of the other listed specialties.

### Data and Statistical Methodology

To enable comparisons of the results, descriptive statistics were calculated as means and standard deviations. For comparisons we avoided any parametric assumptions, by considering the rank correlation. For testing significance, the number of device trials divided by the total number of trials were computed over the study period, and the comparisons were assessed using rank correlation methodology.

## Results

As shown in Table 1 (and in Figure 1), there were 243 PMD trials (179 multi-center, 64 single-center; representing 154 unique medical devices), ranging from 24 to 71 trials per institution; the average number of PMD trials per institution per year was 1.5. Five of the seven institutions initiated fewer than 30 PMD trials during this time frame. The far greater number of concurrent drug and biologic trials compared to PMD trials is noteworthy (Figure 1A and 1B). Concurrent drug and biologic trials were roughly 20 times the number of PMD trials (Table 1).

**Figure 1:**
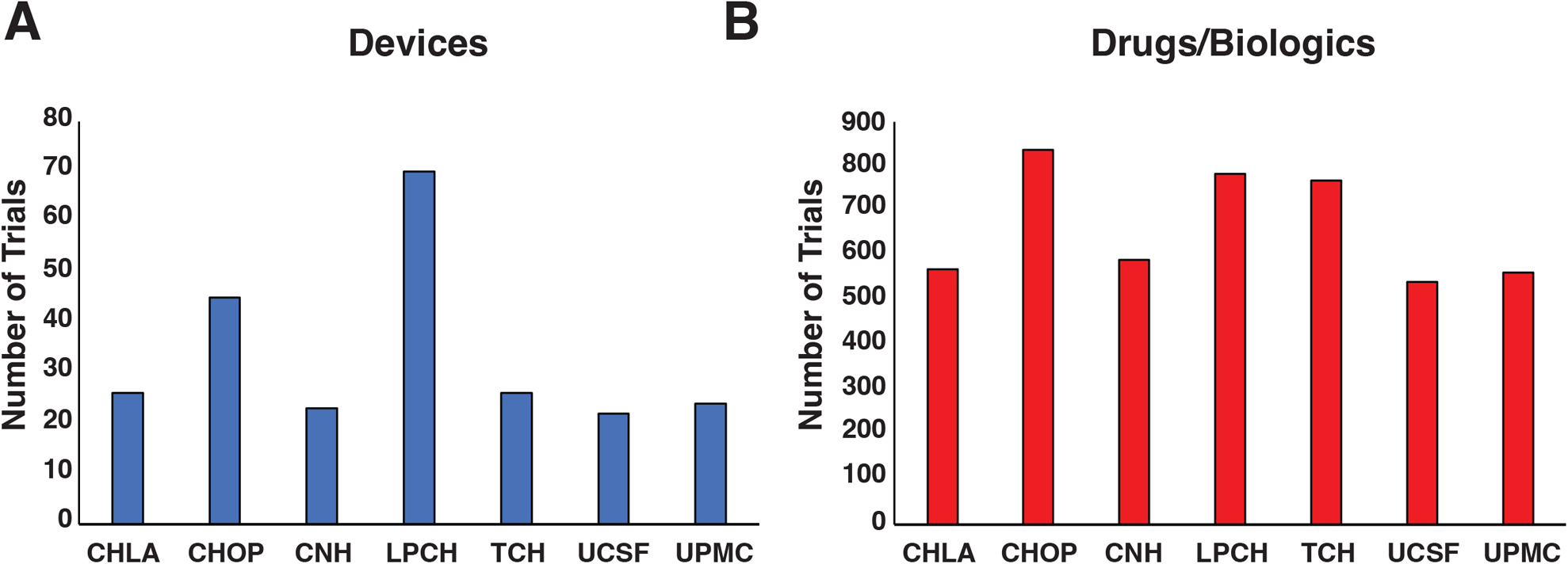
Overview of pediatric medical device (PMD) trials at the member institutions of the FDA PDC Program. **A.** Total PMD trials. **B.** Total concurrent drug and biologic trials between 1999-2022, at each PDC affiliated hospital.

**Table 1:** Clinical Trial Activity at the Pediatric Device Consortia Primary Children’s Hospitals, 1999-2022.

| Institution | Device-Associated Clinical Trial Details |  |  |  |  |  |  |  | Total Device Trials | Total concurrent drug and biologic trials |
| --- | --- | --- | --- | --- | --- | --- | --- | --- | --- | --- |
|  | Completed | Single Sponsored: Industry | Single Sponsored: Other | Multiple Sponsored (NIH co-sponsored) | Multi-center | Class I | Class II | Class III |  |  |
| Children's Hospital Los Angeles | 14 | 8 | 11 | 8 (3) | 18 | 0 | 18 | 9 | 27 | 570 |
| Children's Hospital of Philadelphia | 23 | 14 | 25 | 7 (4) | 30 | 6 | 14 | 26 | 46 | 838 |
| Children's National Hospital | 12 | 6 | 10 | 8 (4) | 15 | 1 | 8 | 15 | 24 | 572 |
| Lucile Packard Children's Hospital Stanford | 48 | 25 | 21 | 25 (19) | 61 | 0 | 45 | 26 | 71 | 818 |
| Texas Children's Hospital | 11 | 13 | 9 | 5 (3) | 22 | 1 | 5 | 21 | 27 | 794 |
| UCSF Benioff Children's Hospital | 15 | 7 | 14 | 2 (0) | 16 | 0 | 11 | 12 | 23 | 550 |
| UPMC Children's Hospital of Pittsburgh | 13 | 10 | 7 | 8 (6) | 17 | 1 | 11 | 13 | 25 | 563 |
| **Total** | **136** | **83** | **97** | **63 (39)** | **179\*** | **9** | **112** | **122** | **243** | **4705** |
| **Mean** | **19.43** | **11.86** | **13.86** | **9.00 (5.57)** | **25.57** | **1.29** | **16.00** | **17.43** | **34.71** | **672.14** |
| **SD** | **13.20** | **6.52** | **6.69** | **7.39 (6.19)** | **16.44** | **2.14** | **13.44** | **6.90** | **17.84** | **135.97** |

*\*These multicenter trials represent 103 unique devices; the single center trials studied 51 unique devices.*

Table 1 shows the number of PMD trials with a single sponsor source: 83 from industry and 97 from other sources (such as academic/research institutions and non-profit organizations). Of the 63 trials that listed multiple funding sources, 39 have the NIH as a co-sponsor, and 14 trials have both NIH and industry sponsorship. There were no examples of NIH sole sponsorship. There were 119 trials that had an industry sponsor, whether as the sole source of sponsorship or in collaboration with other sponsors. In all, industry sponsored studies represented only 2.4% of all combined pediatric clinical trials (Table 1, Total Device Trials, Total concurrent drug and biologic trials). Of the 179 PMD trials that were carried out in multiple institutions, 106 were industry sponsored. Multi-center trials represented 103 unique devices; the single center trials studied 51 unique devices.

Regarding FDA medical device classification, only 9 trials involved Class I devices and the remainder of device trials were comparably distributed between Class II (112 trials) and Class III (122 trials) devices. Of the trials analyzed, two were identified as trials studying combination products, and these trials were *Comparison of Methods of Pulmonary Blood Flow Augmentation in Neonates: Shunt Versus Stent* (The COMPASS Trial) (NCT05268094), involving drug eluting stents, and the *Albuterol Integrated Adherence Monitoring in Children With Asthma* trial (NCT04896645), using a digitally controlled albuterol delivery system.

Additional analysis revealed the inclusion of two trials that involved the use of adult-labeled medical devices. The COMPASS trial is an ongoing prospective multicenter trial, involving up to 300 neonates with ductal dependent congenital heart disease.^22^ The COMPASS trial is investigating efficacy of ductus arteriosus drug eluting stent placement compared to a traditional systemic-to-pulmonary artery surgically created shunt. The drug eluting stents used in this study, sponsored by Healthcore-NERI, represent five different FDA approved stents; all have adult-only labeling. The other clinical trial included in our results that used an adult-labeled device in pediatric subjects involved the Aculaser, a “cold laser” device not intended for pediatric use, that was tested from 2013-2015 in pediatric patients. This trial, sponsored by UCSF, studied children undergoing kidney biopsies, and assessed the Aculaser as a means of acupuncture anesthesia.

Table 2 shows the number of PMD trials by clinical specialty. Cardiovascular and diabetes related devices were the two most common clinical areas with 70 and 63 trials respectively. These results were five-fold higher than the specialties of oncology and otolaryngology, which represented the next highest number of trials. Together, these four specialties were responsible for 64% of all the PMD trials identified. Regarding diabetes trials, it is notable that a single institution, LPCHS, was responsible for 46 of the 63 total trials. The reasons for this predominance are likely multi-factorial, and cannot be determined from the available data.

**Table 2:** Specialized Medical Device Clinical Trial Activity at the Pediatric Device Consortia Primary Children’s Hospitals, 1999-2022.

| **Institution** | **Cardi-**<br>**ova-**<br>**scular** | **Diabe-**<br>**tes** | **Oncol-**<br>**ogy** | **Otolar-**<br>**yngol-**<br>**ogy** | **Ortho-**<br>**pedic** | **Behav-**<br>**ioral** | **Digital** | **Oph-**<br>**tha-**<br>**lmology** | **Infec-**<br>**tious**<br>**Disease** | **Surgery** | **Pulmo-**<br>**nary** | **Neurol-**<br>**ogy/**<br>**neuro-**<br>**surgery** | **Urology** | **Other** |
| --- | --- | --- | --- | --- | --- | --- | --- | --- | --- | --- | --- | --- | --- | --- |
| Children's Hospital Los Angeles | 5 | 7 | 1 | 2 | 3 | 0 | 2 | 0 | 2 | 0 | 0 | 1 | 1 | 3 |
| Children's Hospital of Philadelphia | 14 | 4 | 3 | 2 | 3 | 3 | 4 | 2 | 0 | 1 | 4 | 0 | 0 | 8 |
| Children's National Hospital | 10 | 2 | 3 | 2 | 2 | 1 | 0 | 0 | 1 | 1 | 0 | 1 | 0 | 1 |
| Lucile Packard Children's Hospital Stanford | 12 | 46 | 2 | 3 | 0 | 0 | 0 | 0 | 0 | 2 | 0 | 0 | 1 | 5 |
| Texas Children's Hospital | 13 | 2 | 1 | 1 | 0 | 1 | 0 | 2 | 2 | 1 | 1 | 1 | 0 | 2 |
| UCSF Benioff Children's Hospital | 7 | 1 | 1 | 1 | 2 | 3 | 1 | 0 | 0 | 0 | 0 | 1 | 0 | 5 |
| UPMC Children's Hospital of Pittsburgh | 9 | 1 | 1 | 1 | 0 | 1 | 0 | 3 | 1 | 0 | 0 | 1 | 3 | 4 |
| **Total** | **70** | **63** | **12** | **12** | **10** | **9** | **7** | **7** | **6** | **5** | **5** | **5** | **5** | **28** |
| **Mean** | **10.00** | **9.00** | **1.71** | **1.71** | **1.43** | **1.29** | **1.00** | **1.00** | **0.86** | **0.71** | **0.71** | **0.71** | **0.71** | **4.00** |
| **SD** | **3.27** | **16.45** | **0.95** | **0.76** | **1.40** | **1.25** | **1.53** | **1.29** | **0.90** | **0.76** | **1.50** | **0.49** | **1.11** | **2.31** |

To identify temporal trends in PMD trials, compared to biologic and drug trials, we quantified the number of these trials at five-year increments, with the last time period included (2019-2022) based on 3 ½ years of data (Figure 2). The number of trials initiated for both PMD and drug/biologic trials were relatively constant from 1999 through 2013 (Figure 2A). There was more than a five-fold increase in PMD trials reported during 2014-2018 compared to previous periods. Furthermore, there were 48% more PMD trials from 2019-2022 compared to 2014-2018. PMD trials increased during the full study period, 1999-2022; trials for drugs and biologics showed no significant growth trends (Figure 2A). Figure 2B illustrates the percentage of PMD trials compared to all other trials during these periods. The total number of PMD trials, as a percentage of all pediatric clinical trials, increased, albeit modestly from 1999 to 2013. By 2014, compared to drugs and biologic trials, the percentage of PMD trials begins to significantly (p=0.0083) increase (Figure 2B).

**Figure 2:**
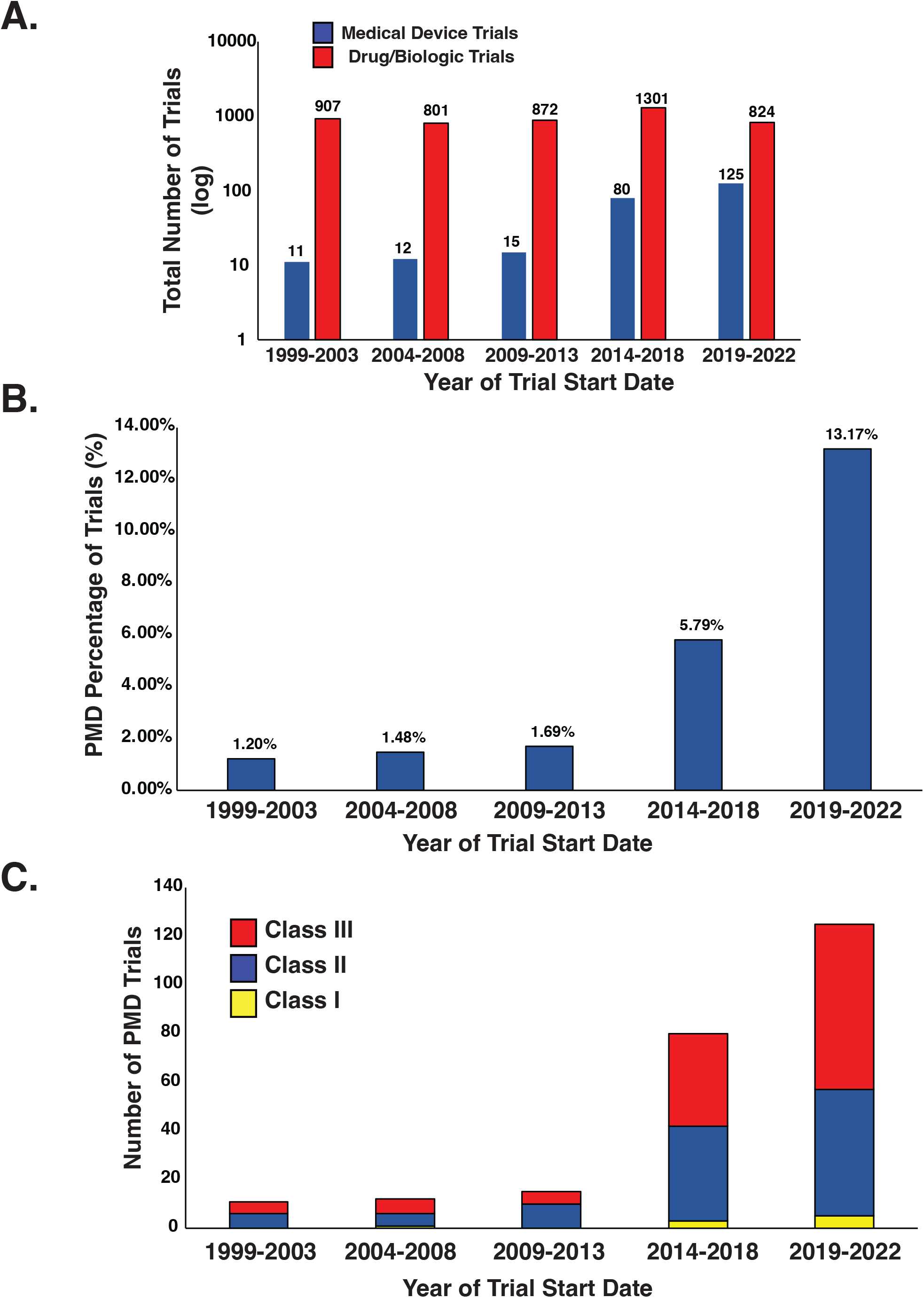
Temporal analysis of PMD trials initiated between 1999 and 2022 at PDC hospitals. **A**. The number of device trials versus drug and biologic trials as a function of trial start date. **B.** Percentage of device trials, compared to total number of trials (*p* = 0.0083). **C.** Breakdown of Device Classification

We also analyzed temporal trends in PMD trials with respect to device classification (Figure 2C). From 2009-2022, the number of trials of Class II devices were 10, 39, and 52 for 2009-2013, 2014-2018, and 2019-2022 respectively. The number of Class III device trials were 5, 38, and 68 for these same respective periods. While most Class I devices are 510(k) exempt and not subject to premarket review requirements like a clinical trial, Class I devices in the trials analyzed in this study demonstrated no sustained increase or definable trend.^19, 23, 24^ Together these data show a progressive and significant increase in PMD trials since 2009. Further analysis of the trials revealed that 6 involved post-market assessment of cardiovascular devices. The initiation of these 6 trials was spread across the timeframe used in this study with 2 post-market trials initiated in 2008; 2 post-approval analysis trials in 2010, 1 trial in 2021; and 1 trial in 2022. These isolated findings did not influence the overall temporal trend in the observed data.

## Discussion

This study profiles the PMD clinical trials performed from 1999-2022 at the cohort of seven academic children’s hospitals primarily affiliated with the FDA PDC grants program. The results show a vast difference between the number of clinical trials involving drugs and biologics compared to PMD, with a roughly 20-fold difference, over the study period, favoring drug trial activity. Specifically, there was an average of 672 trials (Table 1) involving drugs and/or biologics per institution. In contrast, PMD trial activity, while demonstrating a significant increase over time, was relatively modest with a mean of 35 and a range of 23 to 71 PMD trials (Table 1).

For potential insights about current medical device innovation, an informal comparison to adult medical device clinical trials was conducted. We applied search terms used in the present study, with age criteria changed to those for adults, and assessed clinical trial activity for adult medical device development at the 10 top ranked hospitals per U.S. News & World Report’s 2021-2022 Best Hospitals Honor Roll.^25^ The numbers of medical device clinical trials for adults at these institutions ranged from 132-634 per institution over the same time period. This is roughly 6-9 times greater than that of the seven children’s hospitals in our study (Table 1, 23-71 PMD trials per children’s hospital).

The clinical trial infrastructure necessary to support PMD trials has been the subject of relatively few publications,^26, 27^ and has not been broadly defined or characterized; this represents an important issue to be addressed for future PMD trial growth. The average rate of new PMD trial initiation in this study, 1.5 per year (Table 1), likely did not require a unique infrastructure at the children’s hospitals studied, but the availability of such an infrastructure would be beneficial. Many infrastructural elements would support effective, efficient, and optimal growth in PMD trials. Critical needs to support PMD trials include, but are not limited to: IRB personnel and study coordinators with device expertise, engineering resources, clinical trials offices, and device-specific data management services; ideally, all personnel should have relevant expertise in pediatrics. Addressing these needs and the future growth in PMD trial activity reported in this study (Figure 2) may benefit from both institutional and national planning.

The FDA’s PDC grants program was established by Congress in 2007, with grants initiated in 2009. ^16^ Our results in Figure 2 showed that between 2014-2022 there was significant growth in PMD trials at the PDC member institutions. Membership in the PDCs is awarded to academic institutions with an established level of expertise in the PMD field. The increase in PMD trials roughly coinciding with the establishment of the FDA-academic institution partnership, strongly suggests that PMD development and testing could have been responsive to the establishment of FDA support. Thus, the establishment of additional collaborations for pediatric device stakeholders may provide necessary infrastructure to facilitate an increase in PMD trials. Similarly, FDA CDRH initiatives, such as the System of Hospitals for Innovation in Pediatrics-Medical Devices (SHIP-MD), may help advance PMD clinical trials^2^ via establishment of a national coordinated and integrated network; and additionally, de-risk the total product lifecycle for pediatric medical device development, potentially leading to greater investment in and advancement of technology translation for the unique needs of children.

There are limitations of the study design that are noteworthy. The retrospective nature of the methodology involves a selection bias and does not control for changes that likely occurred over the timespan studied. Nevertheless, these data represent a cross-sectional perspective that is useful for understanding PMD clinical trial operations, especially for multi-center studies important for rare disease populations. The use of clinicaltrials.gov as the data source for our studies also has limitations. By way of background, the FDA Modernization Act of 1997 mandated the development of clinicaltrials.gov.^28^ Further provisions in the Final Rule for Clinical Trials Registration and Results Information Submission (42 CFR Part 11) included establishment of a checklist-based tool to assist Responsible Parties in evaluating whether their clinical trial or study is an applicable clinical trial (ACT) for registration with clinicaltrials.gov.^29, 30^ FDA has the authority to issue a Notice of Noncompliance for failure to comply with clinicaltrials.gov requirements.^31^ Thus, in view of these regulations it is likely that our survey of clinical trials performed for Class 2 and Class 3 pediatric medical devices was comprehensive. However, Class 1 medical devices may be approved for use without a requirement for a clinical trial, and thus our Class 1 clinical trials data cannot be viewed as an indicator of growth in PMD. Additionally, it is also important to note that analyses from data acquired from clinicaltrials.gov has been used in prior studies to assess clinical trial activity, ^32–34^ including PMD trial activity,^34^ despite the limitations mentioned above. In addition, concerning this study’s methodology, clinicaltrials.gov search parameters were constrained by its available features. For example, the “Child” age is birth-17 years old. However, the Federal Food, Drug, and Cosmetic Act (FD&C Act) defines pediatric age as birth through 21 years.^17^ Nevertheless, since medical devices intended for adults are generally suitable for patients in the 18-21 years range, the authors do not think the absence of data from this range significantly limits the conclusions.

## Conclusion

Despite a significant increase in PMD trial activity over the 23-year study period, these trials account for only 5% of the total clinical trials at the 7 pediatric medical centers studied. An even smaller portion (2.4%) of PMD trials was sponsored by industry and potentially intended for device development and labeling for pediatrics. The present results suggest that, although the clinical trials growth reflects increased PMD development, the activity observed is relatively small. Nevertheless, accommodation and promotion of PMD clinical trial activity by relevant programs and policies at the institutional and government levels may foster the advancement of PMD to further address unmet needs.

## Data Availability

All data produced in the present study are available upon reasonable request to the authors

## Role of Funder/Sponsor (if any)

The FDA had no role in the design and conduct of this study

## Abbreviations

FDA: Food and Drug Administration
CDRH: Center for Devices and Radiological Health
PMD: Pediatric medical device(s)
PDC: Pediatric Device Consortia
OOPD: Office of Orphan Products Development
CHLA: Children’s Hospital Los Angeles
CHOP: Children’s Hospital of Philadelphia
CNH: Children’s National Hospital
LPCHS: Lucile Packard’s Children’s Hospital Stanford
TCH: Texas Children’s Hospital
UCSF BCH: University of California San Francisco Benioff Children’s Hospital
UPMC CHP: University of Pittsburgh Medical Center Children’s Hospital of Pittsburgh

## Acknowledgment

The authors express their thanks to Ms. Susan Kerns, Children’s Hospital of Philadelphia, for her administrative assistance.

## References

1. Connor EM, Smoyer WE, Davis JM, et al. Meeting the demand for pediatric clinical trials. Sci Transl Med. 2014;6(227):227fs211

2. Espinoza J, Shah P, Nagendra G, Bar-Cohen Y, Richmond F. Pediatric Medical Device Development and Regulation: Current State, Barriers, and Opportunities. Pediatrics. 2022;149(5)

3. Institute of Medicine. Safe Medical Devices for Children. The National Academies Press. 2006

4. Hell AK, Campbell RM, Hefti F. The vertical expandable prosthetic titanium rib implant for the treatment of thoracic insufficiency syndrome associated with congenital and neuromuscular scoliosis in young children. J Pediatr Orthop B. 2005;14(4):287–293

5. Martin MH, Meadows J, McElhinney DB, et al. Safety and Feasibility of Melody Transcatheter Pulmonary Valve Replacement in the Native Right Ventricular Outflow Tract: A Multicenter Pediatric Heart Network Scholar Study. JACC Cardiovasc Interv. 2018;11(16):1642–1650

6. Ramirez N, Flynn JM, Emans JB, et al. Vertical expandable prosthetic titanium rib as treatment of thoracic insufficiency syndrome in spondylocostal dysplasia. J Pediatr Orthop. 2010;30(6):521–526

7. Peiris V, Xu K, Agler HL, et al. Children and Adults With Rare Diseases Need Innovative Medical Devices. J Med Device. 2018;12(3):0347011–0347018

8. Dimitri P, Pignataro V, Lupo M, et al. Medical Device Development for Children and Young People-Reviewing the Challenges and Opportunities. Pharmaceutics. 2021;13(12)

9. Humes HD, Westover AJ. Experience With Pediatric Medical Device Development. Front Pediatr. 2020;8:79.

10. Hwang TJ, Kesselheim AS, Bourgeois FT. Postmarketing trials and pediatric device approvals. Pediatrics. 2014;133(5):e1197–1202

11. Lee SJ, Cho L, Klang E, Wall J, Rensi S, Glicksberg BS. Quantification of US Food and Drug Administration Premarket Approval Statements for High-Risk Medical Devices With Pediatric Age Indications. JAMA Netw Open. 2021;4(6):e2112562

12. Ghandour H, Weiss AJ, Gaudino M, et al. Public reporting for coronary artery bypass graft surgery: The quest for the optimal scorecard. J Thorac Cardiovasc Surg. 2022

13. Jenkins KJ, Beekman RH, Vitale MG, Hennricus WL. Off-Label Use of Medical Devices in Children. Pediatrics. 2017;139(1):1–4

14. Samuels-Reid JH, Blake ED. Pediatric medical devices: a look at significant US legislation to address unmet needs. Expert Rev Med Devices. 2014;11(2):169–174

15. Bleicher EW. Encouraging research and development of pediatric medical devices through legislative and regulatory action: the Pediatric Medical Device Safety and Improvement Act of 2007 in context. Food Drug Law J. 2009;64(3):531–564

16. Ulrich LC, Joseph FD, Lewis DY, Koenig RL. FDA’s pediatric device consortia: national program fosters pediatric medical device development. Pediatrics. 2013;131(5):981–985

17. The US Food and Drug Administration. Pediatric Medical Devices. 2022;Retrieved August 25, 2022(September 22)

18. The US Food and Drug Administration. How to Determine if Your Product is a Medical Device. 2019;Retrieved September 15, 2022

19. The US Food and Drug Administration. Classify Your Medical Device. 2020;Retrieved September 15, 2022

20. The US Food and Drug Administration. Policy for Device Software Functions and Mobile Medical Applications: Guidance for Industry and Food and Drug Administration Staff. 2019(September 27, 2019)

21. The US Food and Drug Administration. What is Digital Health? U.S. Food and Drug Administration. 2020;Retrieved June 3, 2022(September 22)

22. Aggarwal V, Dhillon GS, Penny DJ, Gowda ST, Qureshi AM. Drug-Eluting Stents Compared With Bare Metal Stents for Stenting the Ductus Arteriosus in Infants With Ductal-Dependent Pulmonary Blood Flow. Am J Cardiol. 2019;124(6):952–959

23. The US Food and Drug Administration. Class I and Class II Device Exemptions. 2022(Retrieved September 15, 2022)

24. The National Archives. Part 11 – Clinical Trials Registration and Results Information and Submission. Code of Federal Regulations. 2022

25. Harder B. America’s Best Hospitals: the 2021-22 Honor Roll and Overview. US News and World Report. 2021(July, 27)

26. Bogue C, DiMeglio LA, Maldonado S, et al. Special article: 2014 Pediatric Clinical Trials Forum. Pediatr Res. 2016;79(4):662–669

27. Hirschfeld S, Lagler FB, Kindblom JM. Prerequisites to support high-quality clinical trials in children and young people. Arch Dis Child. 2020

28. The US Food and Drug Administration. Food and Drug Administration Amendments Act of 2007: Public Law 110-85. 2007;Retrieved August 25, 2022

29. ClinicalTrials.gov. Checklist for Evaluating Whether a Clinical Trial or Study is an Applicable Clinical Trial (ACT) Under 42 CFR 11.22(b) for Clinical Trials Initiated on or After January 18, 20171. 2018(accessed January 5, 2023)

30. ClinicalTrials.gov. Frequently Asked Questions. 2022(accessed June 27, 2023)

31. The US Food and Drug Administration. ClinicalTrials.gov - Notices of Noncompliance and Civil Money Penalty Actions. 2022;Retrieved June 27, 2023

32. Califf RM, Zarin DA, Kramer JM, Sherman RE, Aberle LH, Tasneem A. Characteristics of clinical trials registered in ClinicalTrials.gov, 2007-2010. JAMA. 2012;307(17):1838–1847

33. Gresham G, Meinert JL, Gresham AG, Meinert CL. Assessment of Trends in the Design, Accrual, and Completion of Trials Registered in ClinicalTrials.gov by Sponsor Type, 2000-2019. JAMA Netw Open. 2020;3(8):e2014682

34. Hill KD, Chiswell K, Califf RM, Pearson G, Li JS. Characteristics of pediatric cardiovascular clinical trials registered on ClinicalTrials.gov. Am Heart J. 2014;167(6):921–929 e922

